# Implications of the school-household network structure on SARS-CoV-2 transmission under different school reopening strategies in England

**DOI:** 10.1101/2020.08.21.20167965

**Authors:** James D. Munday, Katharine Sherratt, Sophie Meakin, Akira Endo, Carl A B Pearson, Joel Hellewell, Sam Abbott, Nikos I. Bosse, CMMID COVID 1-9 Working Group, Katherine E. Atkins, Jacco Wallinga, W. John Edmunds, Albert Jan van Hoek, Sebastian Funk

**Author notes:** Authors contributed equally (Albert Jan van Hoek, Sebastian Funk). The following authors were part of the Centre for Mathematical Modelling of Infectious Disease COVID-19 working group. Each contributed in processing, cleaning and interpretation of data, interpreted findings, contributed to the manuscript, and approved the work for publication: Rosalind M Eggo, David Simons, Kathleen O’Reilly, Timothy W Russell, Rachel Lowe, Quentin J Leclerc, Jon C Emery, Petra Klepac, Emily S Nightingale, Matthew Quaife, Kevin van Zandvoort, Gwenan M Knight, Thibaut Jombart, C Julian Villabona-Arenas, Eleanor M Rees, Charlie Diamond, Megan Auzenbergs, Graham Medley, Anna M Foss, Georgia R Gore-Langton, Arminder K Deol, Mark Jit, Hamish P Gibbs, Simon R Procter, Alicia Rosello, Christopher I Jarvis, Yang Liu, Rein M G J Houben, Stéphane Hué, Samuel Clifford, Billy J Quilty, Amy Gimma, Damien C Tully, Fiona Yueqian Sun, Kiesha Prem.

## Abstract

**Background:** School closures are a well-established non-pharmaceutical intervention in the event of infectious disease outbreaks, and have been implemented in many countries across the world, including the UK, to slow down the spread of SARS-CoV-2. As governments begin to relax restrictions on public life there is a need to understand the potential impact that reopening schools may have on transmission.

**Methods:** We used data provided by the UK Department for Education to construct a network of English schools, connected through pairs of pupils resident at the same address. We used the network to evaluate the potential for transmission between schools, and for long range propagation across the network, under different reopening scenarios.

**Results:** Amongst the options evaluated we found that reopening only Reception, Year 1 and Year 6 (4-6 and 10-11 year olds) resulted in the lowest risk of transmission between schools, with outbreaks within a single school unlikely to result in outbreaks in adjacent schools in the network. The additional reopening of Years 10 and 12 (14-15 and 16-17 year olds) resulted in an increase in the risk of transmission between schools comparable to reopening all primary school years (4-11 year olds). However, the majority of schools presented low risk of initiating widespread transmission through the school system. Reopening all secondary school years (11-18 year olds) resulted in large potential outbreak clusters putting up to 50% of households connected to schools at risk of infection if sustained transmission within schools was possible.

**Conclusions:** Reopening secondary school years is likely to have a greater impact on community transmission than reopening primary schools in England. Keeping transmission within schools limited is essential for reducing the risk of large outbreaks amongst school-aged children and their household members.

## Introduction

School closures are one of many non-pharmaceutical interventions that can be employed during epidemics of droplet infections, such as influenza, to reduce transmission, and can be highly effective^1–3^. However, there are substantial societal and economic costs associated with closing large numbers of schools, such as limiting children’s access to education and requiring caregivers to stay at home (impacting on household income and on economic activity) ^4–8^, which can affect deprived households most^9^. As with any public health intervention, it is important for policy makers to balance the public health benefits of school closures with the associated economic and social impact. To do this effectively, clear understanding of the relative benefit of closing schools and therefore the potential impact of reopening is required.

School closures were introduced as a central component of the response to the COVID-19 outbreak in many countries around the world^10^. The UK closed all schools on 23^rd^ March 2020 to all but the children of essential workers and the most vulnerable. Schools in England remain closed to the majority of students until the beginning of the coming academic year (September 2020). Although reported cases of COVID-19 are low amongst school-aged children, the role of children in transmitting COVID-19 remains unclear.^11–15^ It remains an open question whether reopening schools might increase transmission within the community and therefore lead to a resurgence of the disease in the population. Notwithstanding the poorly quantified risk, there is growing concern regarding the potential impact of prolonged closures on the wellbeing of the population at large^16^. A recent report from the Royal Society voices concerns that maintaining widespread closures does not just pose a risk to children’s wellbeing in the immediate term but may also have long term consequences for the skill level of the future workforce and therefore economic growth of the UK^17^.

The potential impact of reopening schools on transmission is twofold: firstly, the number of potentially infectious contacts increases through children mixing in schools. Secondly, transmission within schools can facilitate transmission between households, and households with multiple school-aged children may act as a route for transmission between schools. This second impact can be considered as a network of schools and households linked by pupils. While strict stay-at-home orders (so-called lockdowns) as implemented in many countries had the aim of removing the links of network such that chains of infection could not progress beyond individual households, reopening schools has the potential of reconnecting households with each other such that longer chains of infection can arise.

Here, we investigated the potential impact of reopening schools on the connectivity of the school and household network and, consequently, on the potential size of an outbreak among families with school-aged children, under the assumption that children are effective at transmitting the virus. We did so by using a large data set of household addresses of school children in England to quantify probability of transmission through pupils who reside in a common household as the edges on a network of schools. We used this framework to analyse the potential for these links between schools to form large networks of infectious contact and therefore large outbreak clusters within the school-age population and their household members.

## Methods

### Data

Individual level de-identified data of pupils attending state funded schools in England was provided by the UK Department for Education (DfE) under a formal data sharing agreement. The use of this data was also reviewed and approved (Ref: 22476) by the London School of Hygiene & Tropical Medicine Research Ethics Committee. The data includes an entry for each pupil for each institution they attend, Unique Reference Number (URN) for the school, school postcode, pupil’s postcode and pupil’s address, collected between September and December 2019. We combined the student’s postcode and address to assign a household code for each group of pupils that were found to live at the same address, where we assume each individual address operates as a single household for social distancing purposes. Using our generated household code, we were able to estimate the number of unique contacts between each pair of schools. For each pupil we have included only institutions coded as the pupil’s current main school and have excluded pupils listed as boarders (those who are resident at their school during term time). More details of data cleaning are included in the Supplementary Information.

### Reopening Scenarios

Typically, there are 14 school years in the English school system (Figure 1), which each run from September to September. Children enter Reception aged 4 and complete 7 years of primary school leaving year 6 aged 11. They transition to secondary school into year 7 where all pupils are expected to complete 5 years of secondary education (until the age of 16). At this point children are able to leave school or progress to Further Education (FE), which may be in the same institution as other secondary school years or a separate institution offering only FE courses.

**Figure 1.**
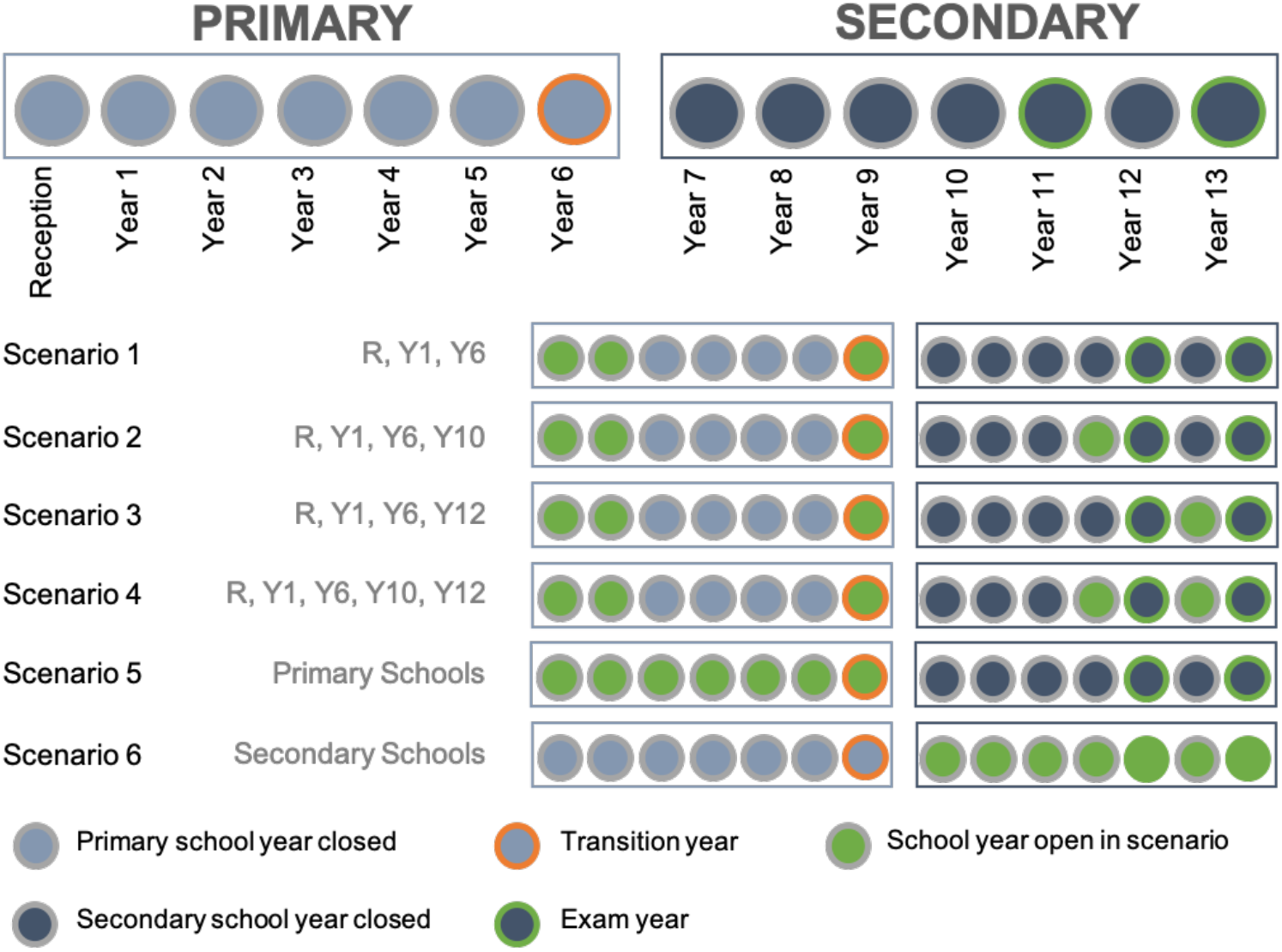
Breakdown of school years in England and reopening scenarios evaluated.

There are exceptions to this two/three institution framework, where some schools offer a different subset of school years (for example the first 3 years of primary education). For this analysis all reopening scenarios are assumed to operate on a school year basis, hence assuming that all children from the appropriate years return regardless of the nature of their institution.

We considered six reopening scenarios relevant to policy in England, illustrated in Figure 1. In each scenario different combinations of year groups return to school: early-years education (Reception and Year 1, i.e. 4-6 year olds) and time-sensitive groups in transition e.g. through exam certifications or transitional years (Years 6, i.e. 10-11 year olds, Year 10, i.e. 14-15 year olds and year 1, i.e. 16-17 year olds)^18^.

### A network of transmission between schools

We used the data to construct a network of schools linked through households. Each edge on the network of schools is weighted by the number of unique contacts between schools that occur through shared households. For example, if in a given household, 2 children attend school *i* and 2 children attend school *j*, this corresponds to 4 unique contacts between school *i* and school *j*. The total number of unique contacts between schools *i* and *j*, denoted by *C_ij_*, is the sum of unique contacts over all households. (Figure 2). Concretely,

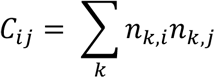

**Figure 2.**
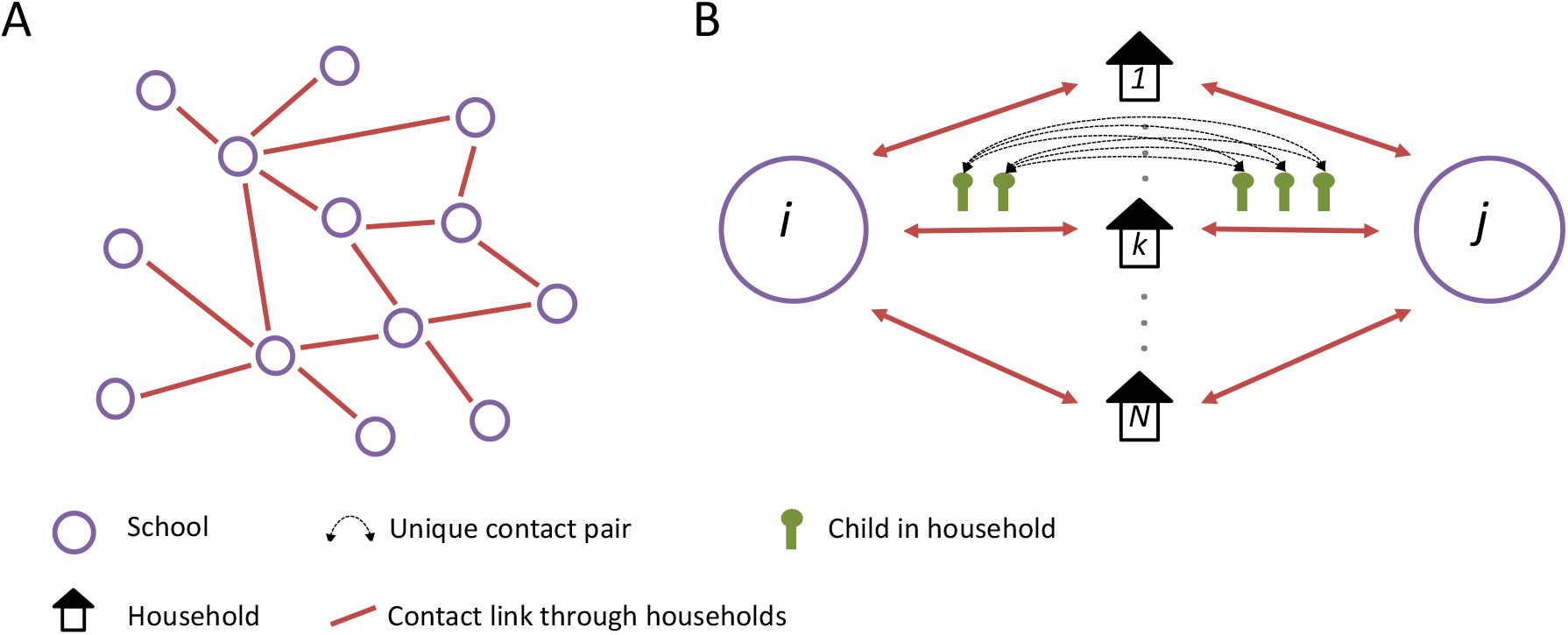
A network of schools linked by households. A) A network of schools constructed such that schools are connected when contact is made between pupils of different schools within a household. B) The strength of contact between schools is quantified by calculating the number of unique contact pairs (one child in each school). The number of pairs per household is the product of the number of children who attend school *i* and the number of children who attend school *j*. The total number of unique pairs is the sum of unique pairs over all, *N*, households, *k*, with children attending both school *i* and *j*.

Where *n_ki_* is the number of children in household *k* who attend school *i*.

From this network, we created a transmission probability network (Figure 3) where we estimated the probability of transmission between schools *i* and *j (P_trans_,_ij_*)

**Figure 3.**
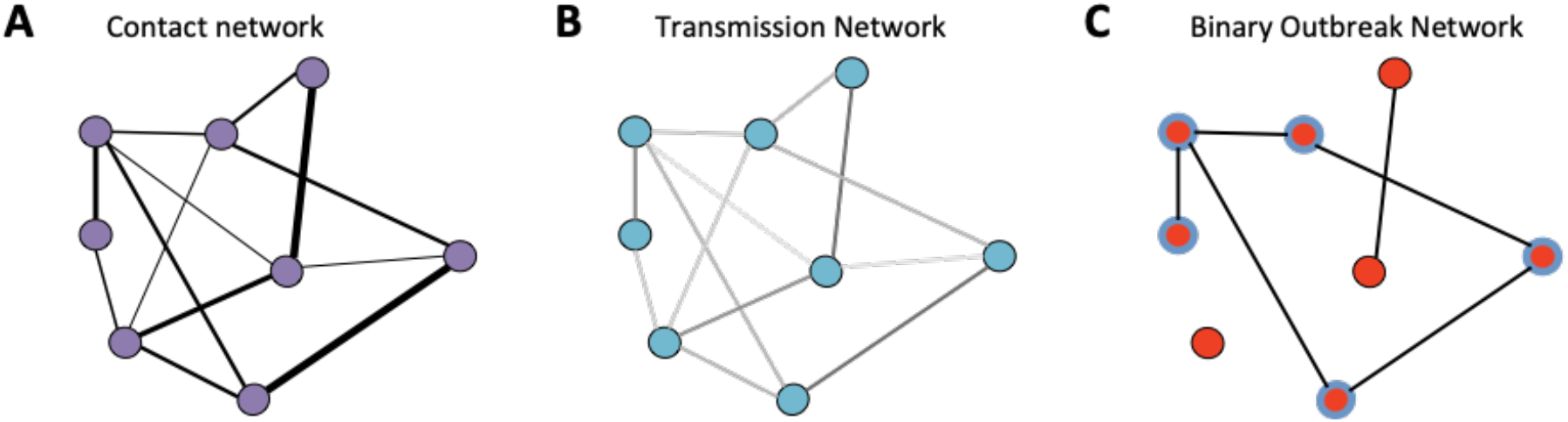
A) A schematic of a contact network, the width of the edges shows the number of unique contact pairs between schools. B) A schematic of a transmission probability network calculated from the contact network; the shading of the edges shows the relative probability of transmission between schools. C) A schematic of a realisation of a binary outbreak network (sampled from B), where edges are weighted 1 with probability given by the equivalent edge in the transmission network, or 0 otherwise. Blue highlighted nodes show those in the largest connected component. In each network nodes show the location of schools.

We defined the definition of transmission between schools as an outbreak in one school leading to an outbreak in an adjacent school on the network. We simplify within-household transmission such that only direct transmission between contact pairs occurs (neglecting the potential for transmission through other members of a household) and hence approximate the transmission probability between schools through a single contact pair as.

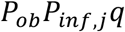

where *P_ob_* is the probability of an outbreak in school *i* given one infection, *P_inf,j_* is the probability of a child in school *j* being infected and *q* is the probability of transmission between children in the same household.

The probability of transmission between schools *j* and *i* through all contact pairs can be approximated as

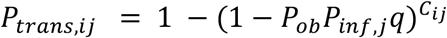

The probability of an outbreak *P_ob_* in any given school with reproduction number *R* was assumed to be,

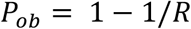

which reflects a geometric distributed contact rate within the school^19^.

We assumed homogeneous mixing within the school population. We then approximated the probability of a student in school *j* being infected *P_j_^I^* based on the expected final size^19^ of an outbreak with reproduction number *R*,

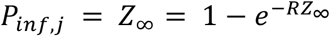

where *Z_∞_* is the final outbreak size as a proportion of the school population.

We set *q*, the per-contact probability of transmission between children in the same household to 0.15 (consistent with estimates of household secondary attack rate of SARS-CoV-2^20^).

We repeated the analysis for a range of *R* values between 1.1 and 1.5, leading to outbreak size between 18 and 58% of school children, broadly spanning the range of reported outbreak sizes of COVID-19 in schools^21,22^.

For each scenario, we assumed all pupils within the years specified attended school and contributed to transmission. We assumed that pupils outside of the specified years did not attend school and therefore did not contribute to transmission. To simulate this condition, we constructed a network using only data of pupils in the specified years.

### Evaluating the network

#### Degree distributions

To summarise how the potential of transmitting to adjacent schools in the network varies with *R* (within school) and the reopening scenario we calculated the distribution of the weighted degree *D* of the transmission network (the distribution of the expected number of schools infected through households by each school) for each scenario, where the weighted degree of school *i, D_i_* was defined as:

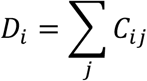

#### Connected components of binary transmission networks

To summarise the potential spread of the virus across the network of schools, we sampled instances of binary outbreak networks, where transmission between each pair of schools either occurs (edge weight of 1) or does not occur (edge weight 0) (Figure 3).

Since transmission probabilities are reciprocal, the eventual number of schools in any outbreak cluster can be defined as a connected component of the outbreak network (i.e. all schools are connected by edges equal to 1). For a particular school *i*, the schools in the same connected component are those that would be infected in an outbreak seeded at that school (*i*). The same schools are those in which a seeded outbreak would eventually infect this school (*i*). Hence the distribution of the connected components gives an indication of expected outbreak size and therefore risk posed to and by individual schools in the network.

Schools vary in size considerably, with large differences between secondary and primary schools. To reflect the size of outbreaks in terms of the number of households at risk, we calculated the number of households with children attending schools within each connected component in the network. Specifically, we calculated the number of unique households with children attending the schools in each component (in the appropriate years for each scenario).

To summarise the risk of larger outbreak clusters, we present the distribution of the number of households associated with each connected component.

## Results

### Network summary

The full contact network (Figure 4) included 21,583 schools, with 4.6 million primary school children and 3.4 million secondary school children in attendance, living at 4.9 million unique addresses. Including only pupils associated with the reopening scenarios reduces the network to containing between 21% and 100% of all schools and between 35% and 66% of all households (Table 1).

**Figure 4.**
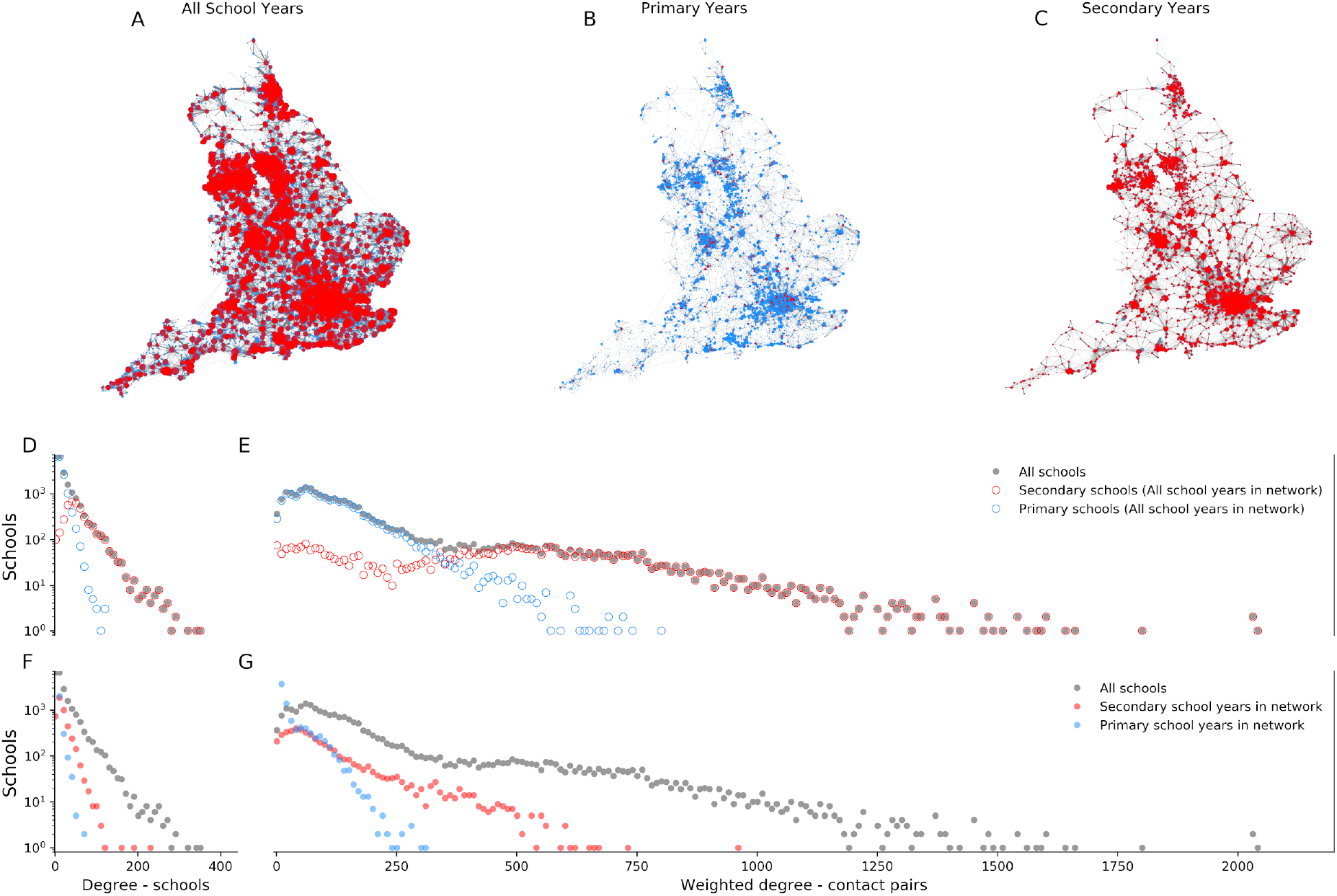
Networks of contact through households between 21,608 state funded schools in England plotted by location. A) Network with all school years in attendance, B) Network with only primary school years in attendance, C) Network with only secondary school years in attendance. Nodes show schools with size determined by the weighted degree of the node (number of unique contact pairs with any other school). Edge widths that indicate the number of unique contact pairs between the schools the edge connects. Red nodes show secondary schools (mean age >= 11 years), blue nodes show primary schools (mean age < 11 years). Followed by degree distributions of the networks of contact through households. D) A histogram of the number of schools connected by at least one contact pair and E) A histogram of the number of unique contact pairs with all other schools in the network including all school years (i.e. that shown in panel A). for all schools (grey) dots, secondary schools (mean age >= 11 years, red circles), and primary schools (mean age < 11 years, blue, circles). F) A histogram of the number of schools connected by at least one contact pair and G) A histogram of the number of unique contact pairs with all other schools in the network including all school years (grey), the network including only secondary school years (blue) and the network including only primary school years (red).

**Table 1.** The number of schools open and households with children attending school in each school reopening scenario.

| Scenario | Number of schools | % of all schools | Number of households | % of all households |
| --- | --- | --- | --- | --- |
| 1\| R, Y1, Y6 | 17,953 | 83 | 1,728,173 | 37 |
| 2\| R, Y1, Y6, Y10 | 21,438 | 99 | 2,211,384 | 45 |
| 3\| R, Y1, Y6, Y12 | 19,982 | 93 | 1,926,090 | 39 |
| 4\| R, Y1, Y6, Y10, Y12 | 21,480 | > 99 | 2,381,729 | 48 |
| 5\| Primary schools (R-Y6) | 17,984 | 83 | 3,267,414 | 66 |
| 6\| Secondary schools (Y7-Y13) | 4,555 | 21 | 2,627,640 | 53 |
| All schools | 21,583 | 100 | 4,927,163 | 100 |

The mean unweighted degree of the schools in the network (average number of schools each school is connected to) was 25 with a maximum of 400. The mean number of contact pairs to all other schools was 184 with a maximum of 2045. (Figure 4). Secondary schools were more connected to the network with higher mean degree, 65 schools, and weighted degree, 480 contact pairs. Primary schools were less connected with mean degree and weighted degrees of 16 schools and 113 contact pairs, respectively and a maximum degree of 127 schools and weighted degree of 806 contact pairs.

When we included only primary school years in the network (i.e. secondary years did not attend school) the mean degree reduced to 6 schools and mean weighted degree to 22 contact pairs. When only secondary school years were included the mean degree and weighted degree reduced to 22 schools and 103 contact pairs respectively.

### Degree distributions of the transmission probability network

With all schools fully open, the mean weighted degree of the transmission probability network (i.e. the mean expected number of schools infected by any individual school) varied between 0.42 for *R* of 1.1, to 3.6 for an *R* of 1.5. The school with the highest weighted degree varied between 4.7 to 35.5 for *R* of 1.1 and 1.5 respectively.

When the network was modified to only include pupils from certain years the mean degrees decreased (Figure 5). Scenario 1 (Reception and years 1 and 6) had the lowest mean weighted degree (0.01 - 0.09) for all values of R, suggesting that on average each school had approximately 1 - 10% chance of infecting one other school. The maximum weighted degree ranged between 0.13 and 1.2, i.e. if an outbreak occurred in the most connected school, it would be expected to infect 1.2 other schools with *R* of 1.5. Scenario 6 (opening secondary schools only) had the highest mean weighted degree, 0.26 - 2.6 across values of *R* 1.1 to 1.5 suggesting that even at low *R* (1.1) there would be approximately a 25% chance, on average, of infecting a second school and at high *R* (1.5) each school would on average infect 2 or 3 schools during an outbreak. After Scenario 6, Scenario 5 (primary schools only) had the highest mean degree, between 0.05 and 0.45. Scenarios 2 - 4, which all combined some partial opening of primary and secondary schools, had relatively similar degree distributions to that of fully opening only, primary schools (Table 2). Of these, Scenario 3 (Reception and years 1, 6 and 12) had the lowest mean degree for each value of *R*, between 0.01 and 0.15.

**Figure 5.**
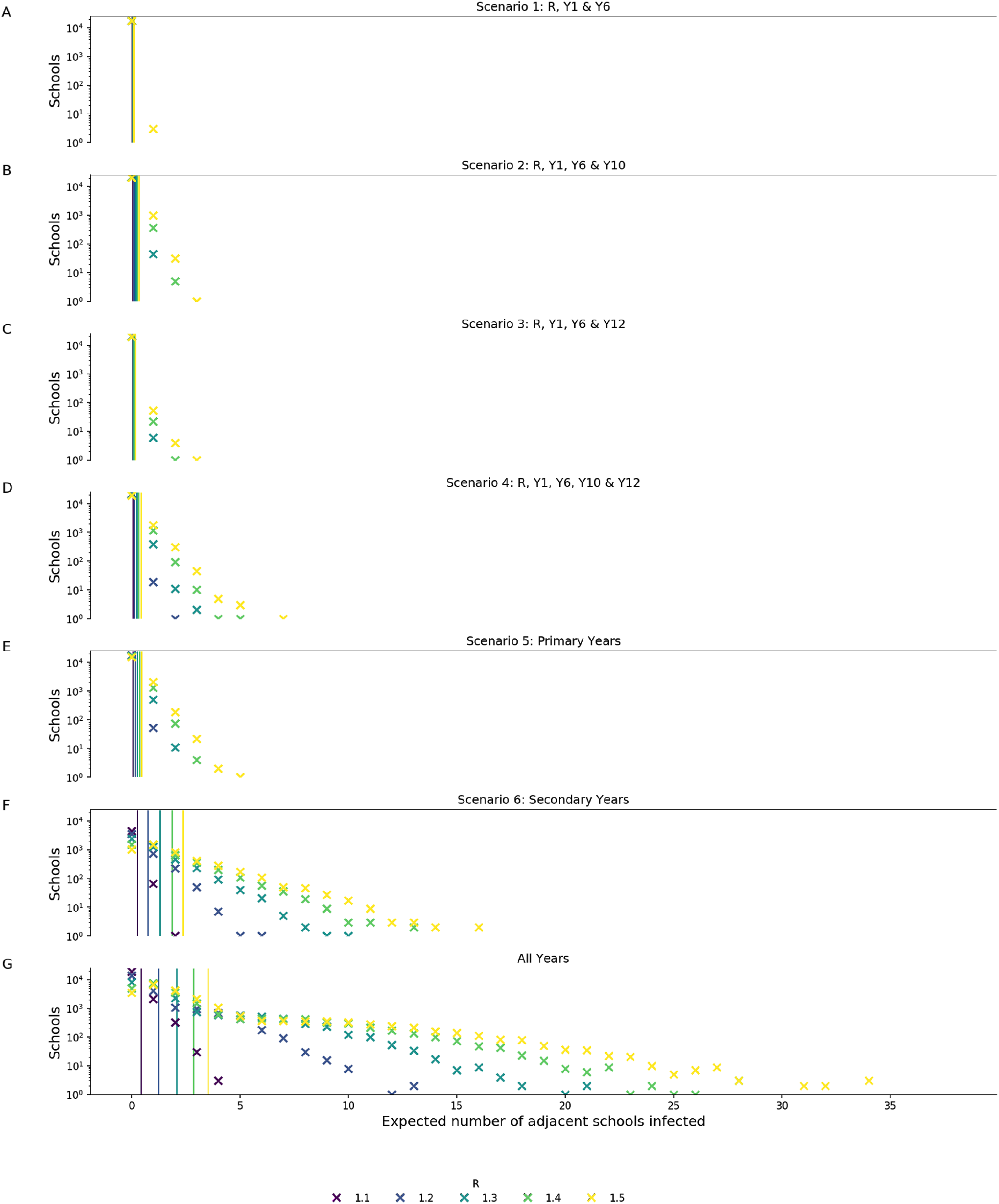
Weighted degree distribution (expected number of schools infected by each school) of the transmission probability network for each of the reopening scenarios considered for R values of 1.1 to 1.5. A - F show scenarios 1 - 6 respectively G shows the network with all school years in attendance. Vertical lines show the mean degree for each value of R.

**Table 2.** Median, mean and maximum weighted degree on the transmission probability network (expected number of schools infected by each school) and median and range of largest component size, households and schools, over 100 realisations of the binary outbreak networks, for each scenario for *R* values between 1.1 and 1.5.

| Scenario | R | Weighted degree of transmission probability network |  |  | Households in largest component of binary outbreak networks |  |  | Schools in largest component binary outbreak networks |  |  |
| --- | --- | --- | --- | --- | --- | --- | --- | --- | --- | --- |
|  |  | Median | Mean | max | median | low | high | median | low | high |
| 1 | 1.1 | 0.00 | 0.01 | 0.13 | 630 | 596 | 889 | 3 | 2 | 4 |
|  | 1.2 | 0.02 | 0.02 | 0.39 | 843 | 672 | 1,154 | 4 | 3 | 5 |
|  | 1.3 | 0.03 | 0.05 | 0.67 | 1,023 | 800 | 1,350 | 5 | 4 | 7 |
|  | 1.4 | 0.04 | 0.07 | 0.95 | 1,313 | 1,000 | 1,893 | 7 | 5 | 9 |
|  | 1.5 | 0.06 | 0.09 | 1.20 | 1,631 | 1,214 | 2,390 | 9 | 7 | 12 |
| 2 | 1.1 | 0.02 | 0.03 | 0.27 | 1,009 | 828 | 1,389 | 5 | 4 | 6 |
|  | 1.2 | 0.06 | 0.10 | 0.87 | 2,154 | 1,569 | 3,778 | 10 | 8 | 17 |
|  | 1.3 | 0.12 | 0.18 | 1.60 | 4,790 | 3,262 | 8,693 | 23 | 17 | 40 |
|  | 1.4 | 0.17 | 0.26 | 2.37 | 12,529 | 7,940 | 21,818 | 66 | 41 | 114 |
|  | 1.5 | 0.23 | 0.34 | 3.11 | 29,517 | 21,151 | 52,983 | 171 | 112 | 272 |
| 3 | 1.1 | 0.01 | 0.01 | 0.30 | 1,166 | 1,166 | 1,408 | 3 | 3 | 5 |
|  | 1.2 | 0.02 | 0.04 | 0.98 | 1,491 | 1,166 | 2,078 | 6 | 5 | 9 |
|  | 1.3 | 0.04 | 0.08 | 1.82 | 2,173 | 1,651 | 3,685 | 10 | 8 | 16 |
|  | 1.4 | 0.07 | 0.11 | 2.71 | 4,047 | 2,686 | 6,355 | 19 | 13 | 32 |
|  | 1.5 | 0.09 | 0.15 | 3.58 | 7,245 | 4,402 | 13,766 | 36 | 22 | 71 |
| 4 | 1.1 | 0.02 | 0.04 | 0.66 | 1,732 | 1,366 | 2,396 | 6 | 5 | 8 |
|  | 1.2 | 0.08 | 0.12 | 2.13 | 4,853 | 3,301 | 7,513 | 18 | 14 | 32 |
|  | 1.3 | 0.14 | 0.22 | 3.91 | 25,024 | 14,540 | 42,580 | 114 | 70 | 186 |
|  | 1.4 | 0.21 | 0.33 | 5.74 | 174,846 | 87,381 | 226,272 | 852 | 427 | 1,139 |
|  | 1.5 | 0.27 | 0.44 | 7.50 | 327,433 | 291,536 | 403,243 | 1,760 | 1,544 | 2,228 |
| 5 | 1.1 | 0.02 | 0.05 | 0.60 | 1,956 | 1,540 | 2,853 | 6 | 4 | 8 |
|  | 1.2 | 0.08 | 0.15 | 1.46 | 5,366 | 3,947 | 9,253 | 16 | 12 | 26 |
|  | 1.3 | 0.14 | 0.26 | 2.61 | 19,639 | 13,524 | 32,166 | 59 | 38 | 90 |
|  | 1.4 | 0.22 | 0.36 | 3.87 | 55,770 | 36,732 | 106,602 | 167 | 105 | 337 |
|  | 1.5 | 0.29 | 0.45 | 5.10 | 126,561 | 76,626 | 229,320 | 418 | 257 | 768 |
| 6 | 1.1 | 0.19 | 0.26 | 2.21 | 44,644 | 26,654 | 96,454 | 50 | 30 | 106 |
|  | 1.2 | 0.60 | 0.81 | 6.46 | 718,224 | 639,961 | 887,222 | 960 | 842 | 1,212 |
|  | 1.3 | 1.08 | 1.43 | 10.60 | 1,588,903 | 1,299,424 | 1,859,976 | 2,341 | 1,916 | 2,758 |
|  | 1.4 | 1.57 | 2.04 | 13.98 | 2,221,866 | 2,098,546 | 2,345,972 | 3,449 | 3,225 | 3,635 |
|  | 1.5 | 2.02 | 2.60 | 16.91 | 2,450,215 | 2,314,264 | 2,502,364 | 3,904 | 3,658 | 3,998 |

### Connected components of binary outbreak networks

The largest connected component of the realisations of the binary outbreak networks, that is, the number of schools in the largest connected part of the network, increased with *R* for each scenario, increasing the number of households at risk (Figure 6, figure S1 (Supplementary Information)).

**Figure 6.**
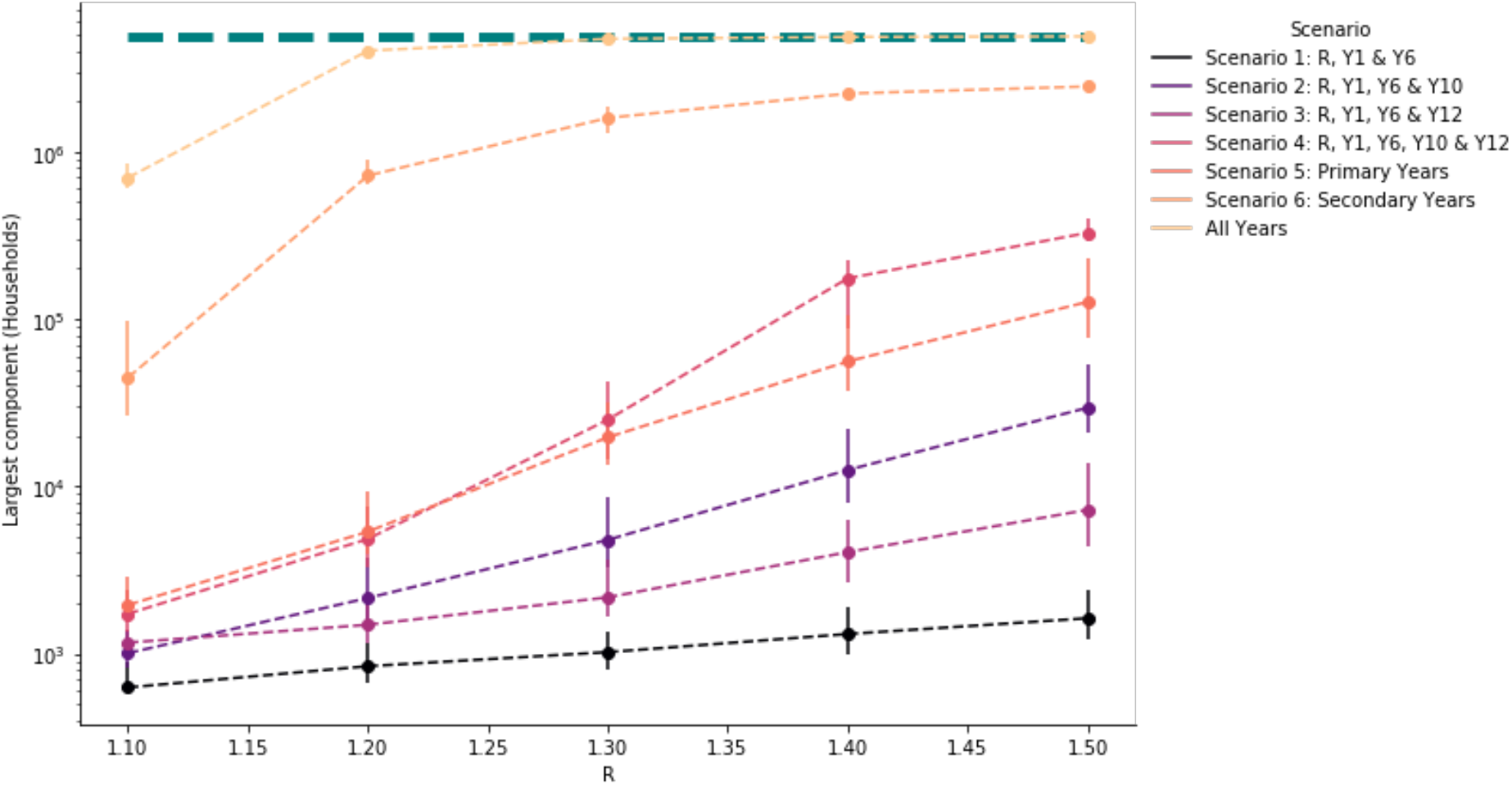
The number of households with children attending a school in each largest connected component of the binary transmission networks (estimated potential outbreak cluster size) generated from transmission probability networks for school reopening scenarios. Vertical bars show the 90% credible intervals. The green dashed line shows the total number of households in the school system.

For Scenario 1 (Reception, Year 1, Year 6) the median largest components simulated ranged between 3 and 9 schools or 630 and 16,031 households across *R* values considered, and there were very few exceeding 10 schools in each realisation (Figure 7), these connected components typically represented fewer than 1,000 households in total.

**Figure 7.**
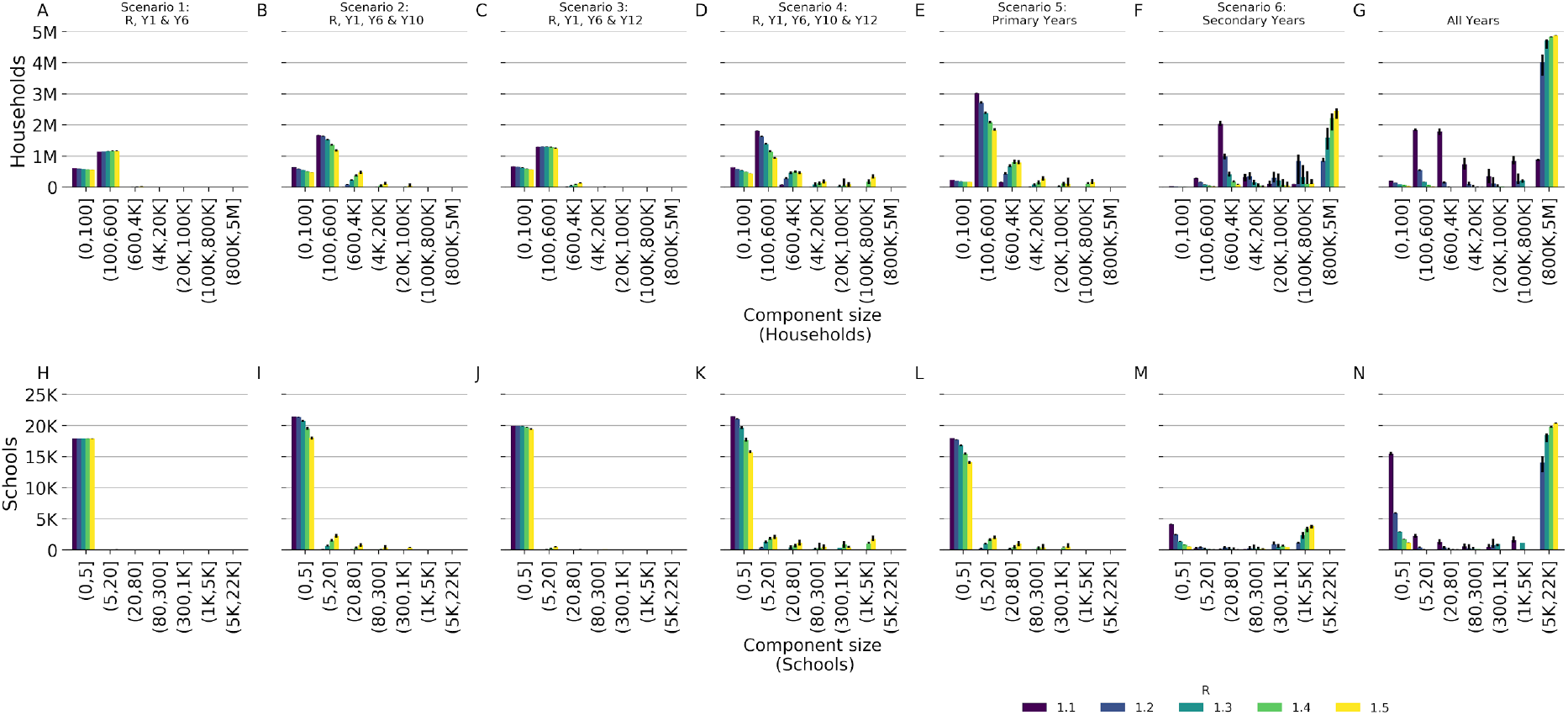
Connected component distribution of the binary outbreak networks generated for school reopening scenarios and *R* values of 1.1 to 1.5 (indicated by colour). By households (A-G), i.e. the number of households in a component size in each bin and by school (H-N). Black bars show 95% credible intervals.

Adding either of secondary school years 10 or 12 to the network (Scenarios 2 and 3) increased the largest connected component size considerably. The size of the largest component was comparable to Scenario 1 at a low *R* of 1.1, with a median largest component size of less than 6 schools for all 5 scenarios. However, the largest connected components for realisations at *R* of 1.5 reached many tens of schools for scenarios 2 and 3 (171 and 36 respectively, compared to 9 for scenario 1) and thousands more households (29,517 and 7,245 compared to 1,631 for scenario 1). Adding both years 10 and 12 had similar largest component size to 1 - 3 at 1.1 (6 schools and 1,732 households) however the largest component at 1.5 was much larger than the other scenarios affecting 1760 schools and 327,433 households. Opening only primary school years (scenario 5) resulted in comparable largest component sizes to scenario 4 at lower values of R but at R of 1.5 resulted in a median largest component of less than a third of schools (median of 418) and less than half as many households (median of 126,561). Largest components were consistently larger when only secondary schools were included in the network, with a median of 50 schools and 44,644 households with an R of 1.1 increasing to 3,904 schools and 2,450,215 households at an R of 1.5 which accounts for 85% of the schools and 93% of households.

Despite the increase in largest component size at higher values of R, for scenarios 1 - 5, the substantial majority of schools remained in small components of less than 5 schools, even with R at 1.5: 17,909 (>99% of schools in the network), 18,024 (84%), 19,442 (97%), 15,716 (73%), 14,130 (79%) for scenarios 1-5 respectively. Whereas for scenario 6, where all secondary school years return, only 538 (12% of schools in the network) schools formed components of less than 5 schools.

## Discussion

Our results suggest that reopening schools with a small selection of school years may only present a small risk of transmission between schools and, consequently, the households of school children. In particular, the analysis highlights the difference in risk posed by secondary schools relative to primary schools, where reopening even a small subset of secondary school years (years 10 and 12) increases the connectivity between schools considerably, whereas opening all primary schools resulted in lower connectivity in the network. Furthermore, opening only secondary schools resulted in the highest connectivity of all the scenarios evaluated.

Recent studies showed that outbreaks in primary schools were smaller than in secondary schools in the same area^21^ and that older children might pose a greater risk of onwards transmission in households than younger ones^23^. In combination, these studies suggest that primary schools pose a lower risk of increasing community infections than secondary schools and support the prioritisation of primary schools for reopening^16^ although if children in secondary schools were better able to practice physical distancing than primary schools, this could act to counterbalance the additional risk. Under the assumption that primary school children transmit the virus less efficiently than older school children^15^, the difference between the scenarios of reopening either primary or secondary schools would be expected to be greater than what we found. In the extreme case where primary school children were not able to transmit the virus at all, the scenario of reopening all years would be the same as reopening only secondary schools. Our assumption is that transmission in school aged children is sufficient to sustain an outbreak within a school i.e. R > 1. Although there is some evidence of transmission within schools^21,22^ and that closing schools reduced the growth rate of the epidemic^24^, other studies have shown that transmission in schools did not contribute greatly to the overall epidemic prior to closure^25,26^. Scientific consensus on this matter remains elusive^27^, and our results should therefore be considered in light of the most recent available evidence to the reader.

Although we found that varying the reproduction number within the schools, *R*, had a substantial impact on the number of households in the largest potential outbreak cluster (indicated by the largest component), there was little impact on the results for the vast majority of schools’ component sizes, suggesting that particular parts of the network were more closely connected than the rest of the network. This could result in particular areas being disproportionately affected following the reopening of schools. Increasing R also had some impact on the weighted degree distribution of the transmission probability network, suggesting that in that case the virus may spread more effectively across connected components even if the eventual outbreak cluster size remained similar. This may impact the effectiveness of targeted interventions, as identifying a school outbreak before an outbreak in an adjacent school has been seeded may become more challenging. This is analogous to challenges in contact tracing due to pre-symptomatic infection^28,29^.

Our network focusses on transmission in schools and households between school-aged children and aims to provide insight into the capacity for transmission within schools and households to develop into large outbreak clusters involving multiple schools. Further, we cannot account for mixing among children from different schools or households occurring outside of school contexts^12^. The data from which the network was constructed, included only state-funded schools in England with children coded as school years Reception to Year 13 in official data, which accounts for 75% of children in this age bracket. The addition of independent schools would increase the size and possibly the connectivity of the network, however only 7% of children in England attend an independent school so the impact may be marginal.

The way we quantified the probability of transmission between schools assumed that each school outbreak reached its unmitigated final size - this may be unlikely before interventions, such as targeted school or class closure are introduced. For example, closure of schools when a small number of cases are reported could be an effective means to curb transmission ^30,31^ early on, however, to the knowledge of the authors, the effectiveness of such reactive closures is yet to be quantified in the context of SARS-COV-2. This framework also implies a well-mixed contact network within each school, final sizes are likely to be smaller if schools implement social bubbles to introduce community structure in the contact network and therefore reduce the probability of a school wide outbreak^32^. The reproduction number was assumed to be invariant between schools, this approach was chosen to maintain the parsimony of the approach, as modelling internal transmission dynamics of individual schools would increase complexity considerably.

We assumed transmission between members of the same household to occur with probability *q =* 0.15, which is consistent with estimates of the household secondary attack rate^20,33^. To assess the robustness of the results to this assumption, we re-ran the analysis with *q =* 0.3 and *q =* 0.08 (supplementary material), where although the sizes of the connected components changed, the relative impact of scenarios remained comparable to the main analysis.

Our analysis provides insight into the potential for school-based and household-based contacts between children to combine to create long chains of transmission which could result in infections within many thousands of households. We highlight that the impact of restarting school on transmission varies substantially between the tested scenarios. Reintroducing primary school years had much lower risk of transmission between schools than secondary school years. We also highlight that maintaining restrictions on contact between children within schools to ensure a low within-school reproduction number may be highly influential, as the rate of transmission between schools increases rapidly with *R* on some parts of the network. Furthermore, such restrictions will be essential for suppressing transmission in the event that all secondary schools are opened. Further analysis using this network may provide more precise guidance, particularly on reactive school closure strategies in the event of detecting a school outbreak, where the network itself may serve as a tool to aid targeted interventions.

Our results are directly applicable to the school system in England. Although the network properties of school systems around the world may vary, we anticipate these results would be qualitatively similar in other settings with broadly comparable education frameworks.

## Data Availability

The data used for this analysis is sensitive and therefore not shareable.

## Authors Contributions

The study was conceived by all of the authors, and designed by JDM, SRM, KS, WJE, AJvH and SFunk. The data was cleaned and matched by JDM and KS. The general “school network” methodology was developed by JDM, KEA, JW and AJvH. The analysis in this study was carried out by JDM. The results were interpreted and the manuscript was written by JDM, SRM, KS and SFunk and edited by all authors.

## Acknowledgements

The authors would like to thank the Her Majesty’s Government’s Department for Education for extracting and arranging access to the appropriate pupil data.

